# State minimum wage, paid sick leave, and food insufficiency during the COVID-19 pandemic

**DOI:** 10.1101/2021.03.01.21252723

**Authors:** Julia Raifman, Elaine Nsoesie, Lorraine T. Dean, Katherine Gutierrez, Will Raderman, Alexandra Skinner, Paul Shafer

## Abstract

**Introduction:** People in low-income households face a disproportionate burden of health and economic consequences brought on by the COVID-19 pandemic, including COVID-19 and food insufficiency. State minimum wage and paid sick leave policies may affect whether people are vulnerable to employment and health shocks to income and affect food insufficiency.

**Methods:** We evaluated the relationship between state minimum wage policies and the outcome of household food insufficiency among participants younger than 65 during the COVID-19 pandemic. We used data from biweekly, state representative Census Pulse surveys conducted between August 19 and December 21, 2020. We conducted analyses in the full population under age 65 years, who are most likely to work, and in households with children. The primary exposure was state minimum wage policies in four categories: less than $8.00, $8.00 to $9.99, $10.00 to $11.99, and $12.00 or more. A secondary exposure was missing work due to COVID-19, interacted with whether participants reported not having paid sick leave. Food insufficiency was defined as sometimes or often not having enough to eat in the past seven days. Very low child food sufficiency was defined as children sometimes or often not eating enough in the past seven days because of inability to afford food. We conducted a multivariable modified Poisson regression analysis to estimate adjusted prevalence ratios and marginal effects. We clustered standard errors by state. To adjust for state health and social programs, we adjusted for health insurance and receipt of supplemental nutrition assistance program benefits, unemployment insurance, and stimulus payments as well as for population demographic characteristics associated with food insufficiency. We conducted subgroup analyses among populations most likely to be affected by minimum wage policies: Participants who reported any work in the past seven days, who reported <$75,000 in 2019 household income, or who had a high school education or less. We conducted falsification tests among participants less likely to be directly affected by policies, ≥65 years or with >$75,000 in 2019 household income.

**Results:** In states with a minimum wage of less than $8.00, 14.3% of participants under age 65 and 16.6% of participants in households with children reported household food insufficiency, while 10.3% of participants reported very low child food sufficiency. A state minimum wage of $12 or more per hour was associated with a 1.83 percentage point reduction in the proportion of households reporting food insufficiency relative to a minimum wage of less than $8.00 per hour (95% CI: −2.67 to −0.99 percentage points). In households with children, a state minimum wage of $12 or more per hour was associated with a 2.13 percentage point reduction in household food insufficiency (95% CI: −3.25 to −1.00 percentage points) and in very low child food sufficiency (−1.16 percentage points, 95% CI: −1.69 to −0.63 percentage points) relative to a state minimum wage of less than $8.00 per hour. Minimum wages of $8.00 to $9.99 and $10.00 to $11.99 were not associated with changes in child food insufficiency or very low child food sufficiency relative to less than $8.00 per hour. Subgroup analyses and sensitivity analyses were consistent with the main results. Estimates were of a lesser magnitude (<0.6 percentage points) in populations that should be less directly affected by state minimum wage policies. Missing work due to COVID-19 without paid sick leave was associated with a 5.72 percentage point increase in the proportion of households reporting food insufficiency (95% CI: 3.59 to 7.85 percentage points).

**Discussion:** Food insufficiency is high in all households and even more so in households with children during the COVID-19 pandemic. Living in a state with at least a $12 minimum wage was associated with a decrease in the proportion of people reporting food insufficiency during the COVID-19 pandemic. Not having paid leave was associated with increases in food insufficiency among people who reported missing work due to COVID-19 illness. Policymakers may wish to consider raising the minimum wage and paid sick leave as approaches to reducing food insufficiency during and after the COVID-19 pandemic.

## Introduction

People, especially children, in low-income households have borne the brunt of the health and economic consequences of the COVID-19 pandemic. As more than 50 million people lost work,^1^ unemployment was concentrated and prolonged for people in low-income households.^2^ More than 20 million people have not had enough money to meet their most basic needs.^3^ There are increases in both food insecurity – typically measured on a ten item scale over a one year period and defined by the US Department of Agriculture (USDA) as households “unable to acquire adequate food for one or more household members because the households had insufficient money and other resources for food” – and food insufficiency – typically measured over 7 days, and defined as when “households sometimes or often did not have enough to eat.^4^” Food insecurity is estimated to have exceeded 20% in the full population and 30% in households with children.^5,6^ Structural racism has created large racial disparities in wealth^7^ and vulnerability to economic shocks, compounded by racial disparities in unemployment;^8^ there are large racial and ethnic disparities in food insufficiency.^9^ There are likely to be significant short and long-run consequences for health and human capital.^10,11^ Already, people in low-income households have reported the highest levels of mental distress^12,13^ and suicidality^14^ during the COVID-19 pandemic, and participation in education declined most for children in low-income households.^2^ The financial costs will likely far exceed the pre-pandemic estimated cost of food insecurity of $167 billion annually.^15,16^

While people in low-income households faced food and housing insecurity, those who could continue working often faced high exposure to COVID-19 through their jobs and work commutes. One estimate found that less than 25% of people with a high school or less education could work from home, whereas 75% of those with post-graduate education could work from home.^17^ Disparities in COVID-19 death rates may reflect COVID-19 exposure at work, with people with less than a high school education 5.3 times and high school graduates 3.4 times more likely to die of COVID-19 relative to people with a postgraduate degree by January 31, 2021.^18^ There were also large racial and ethnic disparities in excess deaths, reaching 53% for Latinx people and more than 30% for Black, Asian, and Native American people, relative to 12% for White people^19^ which may be due to exposure to COVID-19 through work.^20,21^ Essential workers who were more likely to be exposed to and contract COVID-19 may have lost wages due to their illness, furthering economic precarity and food insufficiency. While Congress required large businesses and incentivized smaller businesses to provide to a two-week paid leave in April 2020, more than 100 million workers were exempt from required paid leave, and it expired after December 2020.^22^

The Supplemental Nutrition Assistance Program (SNAP) aims to address food insecurity by providing electronic debit cards that can be spent on food, but in an amount that is often insufficient to cover all food needs in a non-pandemic context.^23^ Increased food prices and food stock disruptions during the COVID-19 pandemic^24^ may have further reduced food affordability and availability for people with SNAP benefits in 2020 and 2021. During the pandemic, Congress allowed states to extend the duration of eligibility and provide the maximum amount to all participants. Congress later increased monthly SNAP benefits by 15% from January through June 2021.^25^ While we did not identify studies on the impact of SNAP during the pandemic, there is evidence that the Pandemic EBT program for children played an important role in reducing food insufficiency.^26^ While SNAP is designed to quickly provide benefits to people who newly qualify based on assets and income,^6^ qualifying and enrolling remain challenging.^27^ In 2017, enrollment ranged from 52% to 100% of eligible people across states.

Other policies that shape funds available to people in low-income households may also affect food insufficiency given that SNAP alone is typically not enough.^23^ Multiple studies indicate that unemployment insurance coverage^28,29^ and amount^29,30^ were associated with reduced food insufficiency during the COVID-19 pandemic.

State minimum wage and paid leave policies may also play an important role in whether people have enough money to live and their vulnerability to income shocks brought on by the COVID-19 crisis.^31^ Higher minimum wages have been associated with lower food insecurity in international comparisons.^32^ There are also several studies indicating that higher minimum wage is associated with reduced suicide^33,34^ and that people in low-income households are at elevated risk of suicide due to shocks that may be brought on by COVID-19 economic conditions, such as unemployment and eviction.^35–38^

Evaluating the relationship between policies and health during the COVID-19 pandemic can inform policies that improve health during the continued pandemic and during the period of recovery in the wake of the pandemic. In this analysis, we evaluated the relationship between state minimum wage policies and food insufficiency. A second objective was to evaluate the relationship between missing work due to COVID-19 infection, paid sick leave, and food insufficiency.

## Methods

### Sample

We used repeated cross sectional Census Pulse survey data representative of individuals in all 50 US states. We used data from Phases 2 and 3, waves 13 to 21, conducted biweekly from August 19 to December 21, 2020 (Phase 1 had a different sampling strategy; in Phases 2 and 3, there were not repeated panel observations of the same participants). We restricted the analysis to people of working age (<65 years) as the population that would be directly affected by minimum wage increases. We also conducted analyses among households with children given excess rates of food insufficiency. We excluded participants who did not respond to questions about food insufficiency or covariates included in the main analysis. We conducted subgroup analyses by race and ethnicity and among those who would be most likely to be affected by increases in minimum wages: 1) People who reported working in the past seven days, 2) people with 2019 household income of less than $75,000, and 3) people with educational attainment of high school or less. We conducted falsification tests among those unlikely to be directly affected by minimum wages, people 65 years and older with household income of $75,000 or more.

### Exposure

The main exposure of interest was state minimum wage policies documented in the COVID-19 US State Policy Database,^39^ which is based on government websites and compiled by two independent research assistant coders to ensure accuracy. We confirmed minimum wage values through comparison with Federal Reserve Economic Data® (FRED) from the St. Louis Federal Reserve Bank.^40^ For several states that exempted small businesses or people in training from minimum wage policies, we used the minimum wage for non-exempt businesses or workers. Nevada has a higher minimum wage for workers who do and do not receive health insurance through their work. As most low-income workers did not have employer-sponsored health insurance, we used the higher minimum wage, for workers without health insurance.. These decisions make our estimates conservative as they reflect the highest minimum wage applicable in a state. In two states that changed the minimum wage during the study period (Connecticut in September and Rhode Island in October), we coded the minimum wage as increasing in the subsequent study wave to allow workers to receive increased paychecks.

We coded the state minimum wage levels into four categories: less than $8.00, $8.00 to $9.99, $10.00 to $11.99, or $12.00 or more per hour (none of the 50 states had a minimum wage of $15 per hour). We chose larger cut points than $1 increments so that there were several (8 or more) states with different geographic locations and other characteristics in each group. We investigated $1 cut points in a sensitivity analysis.

As a secondary objective, we evaluated another exposure potentially related to food insufficiency, a binary indicator for whether participants missed work in the past 7 days because they had COVID-19. We interacted this term with a binary indicator for not having paid sick leave or receiving full or partial pay in response to the question “Are you receiving pay for time not working?”

### Outcomes

The main outcome of interest was a binary indicator of household food insufficiency based on the question, “In the last 7 days, which of these statements best describes the food eaten in your household?” We coded responses of “Sometimes not enough to eat” and “Often not enough to eat” as equal to one and responses of “Enough of the kinds of food (I/we) wanted to eat” and “Enough, but not always the kinds of food (I/we) wanted to eat” as equal to zero.

We also conducted analyses of household food insufficiency and very low child food sufficiency in households with children. In households with children, household food insufficiency was defined in the same way as in the full sample of adults under the age of 65 years. The outcome of very low child food sufficiency was based on the following question, which was only asked of those reporting household food insufficiency, “Please indicate whether the next statement was often true, sometimes true, or never true in the last 7 days for the children living in your household who are under 18 years old. ‘The children were not eating enough because we just couldn’t afford enough food.’” In keeping with prior research,^26^ we coded very low child food sufficiency as equal to one if participants reported “Sometimes true” or “Often true” in response to this question. We coded very low child food sufficiency as zero if household food insufficiency was equal to 0 or if participants responded “Never true” to this question.

### Covariates

We adjusted for several covariates to address state population characteristics and state health and social policies that may confound the relationship between state minimum wage and household food insufficiency. We adjusted for participant-level characteristics, including race and ethnicity (non-Hispanic White, non-Hispanic Black, Non-Hispanic Asian, Non-Hispanic mixed race or another race, or Latinx; the Census Pulse survey does not report whether people are Native American) to account for structural racism shaping disparities in wealth and economic precarity, age group (18 to 24, 25 to 39, 40 to 54, 55 to 64 years), gender identity (woman or man, termed female or male in the Census Pulse question), marital status (married or unmarried), educational attainment (less than high school, high school graduate or equivalent, some college or associate’s degree, or a college graduate). We adjusted for household composition (1, 2, 3, or 4 or more adults and 1, 2, or 3 or more kids). We did not adjust for sexual orientation or sex at birth because the Census Pulse surveys do not collect this information. We also adjusted for work status and other sources of food as binary variables, including whether households reported having lost work during the COVID-19 pandemic, whether they worked in the past 7 days, whether they received unemployment insurance, whether they received free food, and whether they received SNAP benefits. We adjusted for health insurance coverage (public, private, uninsured, or undetermined). We also adjusted for each survey wave. We did not adjust for income group as the main mediator of the relationship between state minimum wage policies and household food insufficiency.

### Analyses

We described the demographic and socioeconomic characteristics of the sample by state minimum wage category. We described household food insufficiency rates and very low child food sufficiency by state minimum wage category. We also described the cumulative proportion of people reporting missing work due to COVID-19 illness by minimum wage.

In the main analysis, we estimated the relationship between state minimum wage category and the proportion of the population under the age of 65 years reporting household food insufficiency. We also estimated household food insufficiency and very low child food sufficiency among participants under the age of 65 years living with children. We estimated adjusted prevalence ratios (aPR) using a multivariable modified Poisson regression analysis to approximate a log binomial regression model^41,42^ and reported marginal effects at the mean values of covariates. We clustered standard errors by state to account for evaluating a state-level policy using repeated observations by state.^43^ We used household survey weights because the outcome was measured at the household level. While the Census Bureau recommends using balanced repeated replication weights, clustering by repeated observations is a more conservative approach to standard error estimation that avoids underestimating standard errors, as noted by other researchers.^28^

We estimated the relationship between state minimum wage category and household food insufficiency among several subgroups that would be more directly affected by minimum wage policies: 1) people who reported working in the past week, 2) people living in households earning less than $75,000 in 2019, and 3) people with a high school diploma or less education. We evaluated whether the relationship between minimum wage and household food insufficiency differed by race to inform how minimum wage policies may affect racial and ethnic disparities in household food insufficiency.^44^ We also conducted falsification tests among people who should not have been as directly affected by minimum wage, including people 65 years and older and people with a 2019 household income of $75,000 or more.

We also conducted several sensitivity analyses. We repeated the main analysis with the minimum wage exposure variable coded as $1.00 increments rather than $2.00 increments. We repeated the main analysis with the outcome of household food insufficiency redefined as food insufficiency due to not being able to pay for food, coded as a binary outcome with a value of one only if participants reported not having enough money to pay for food as the reason for food insufficiency. We also report results for a model without survey weights^45^ and for a model using balanced repeated replication^46^ to estimate standard errors rather than clustering standard errors by state.

## Results

There were 573,215 Census Pulse participants under the age of 65 years during the study period. We excluded 56,102 (9.8%) of participants who did not respond to the question about food sufficiency. We excluded another 9,191 (1.6%) who did not respond to questions about covariates. The study sample consisted of 507,922 participants under the age of 65 years, 229,833 participants in households with children, and 299,107 participants in households with children that reported on child food sufficiency (Table 1). Participants living in states with a minimum wage of less than $8 per hour were more likely to report household income of less than $50,000 in 2019 (33.8% weighted) relative to participants in states with a minimum wage of $12 or more per hour (28.6%). Reports of missing work due to COVID-19 ranged from 1.1% in states with a minimum wage of $12 or more per hour to 1.4% in states with a minimum wage of less than $8.00 per hour. A greater proportion of participants were non-Hispanic Black (15.8%) and a lower proportion were non-Hispanic Asian (2.9%) in states with minimum wage of less than $8 per hour relative to states with a minimum wage of $12 or more per hour (non-Hispanic Black: 5.5% and non-Hispanic Asian: 9.4%).

**Table 1:**
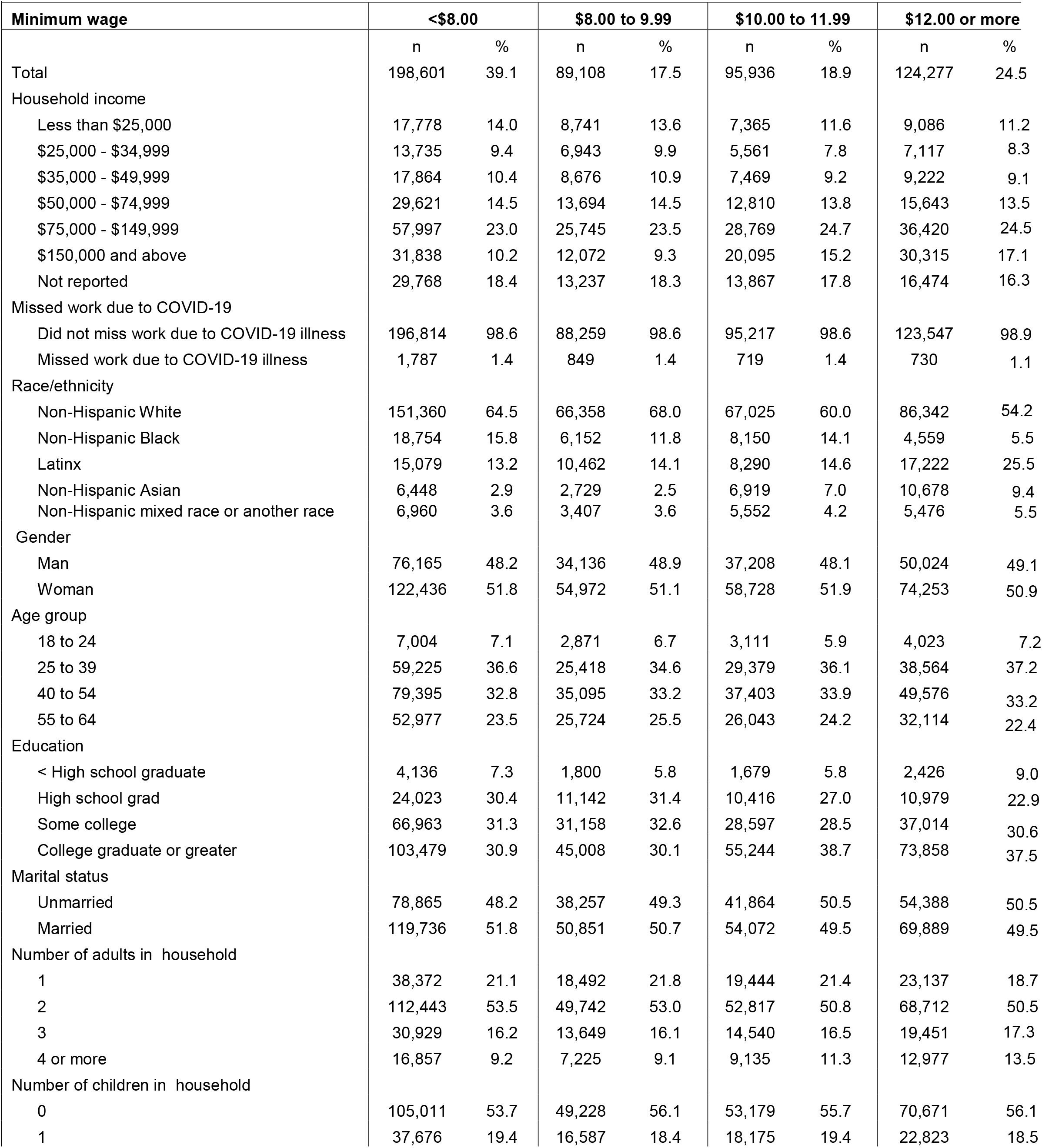
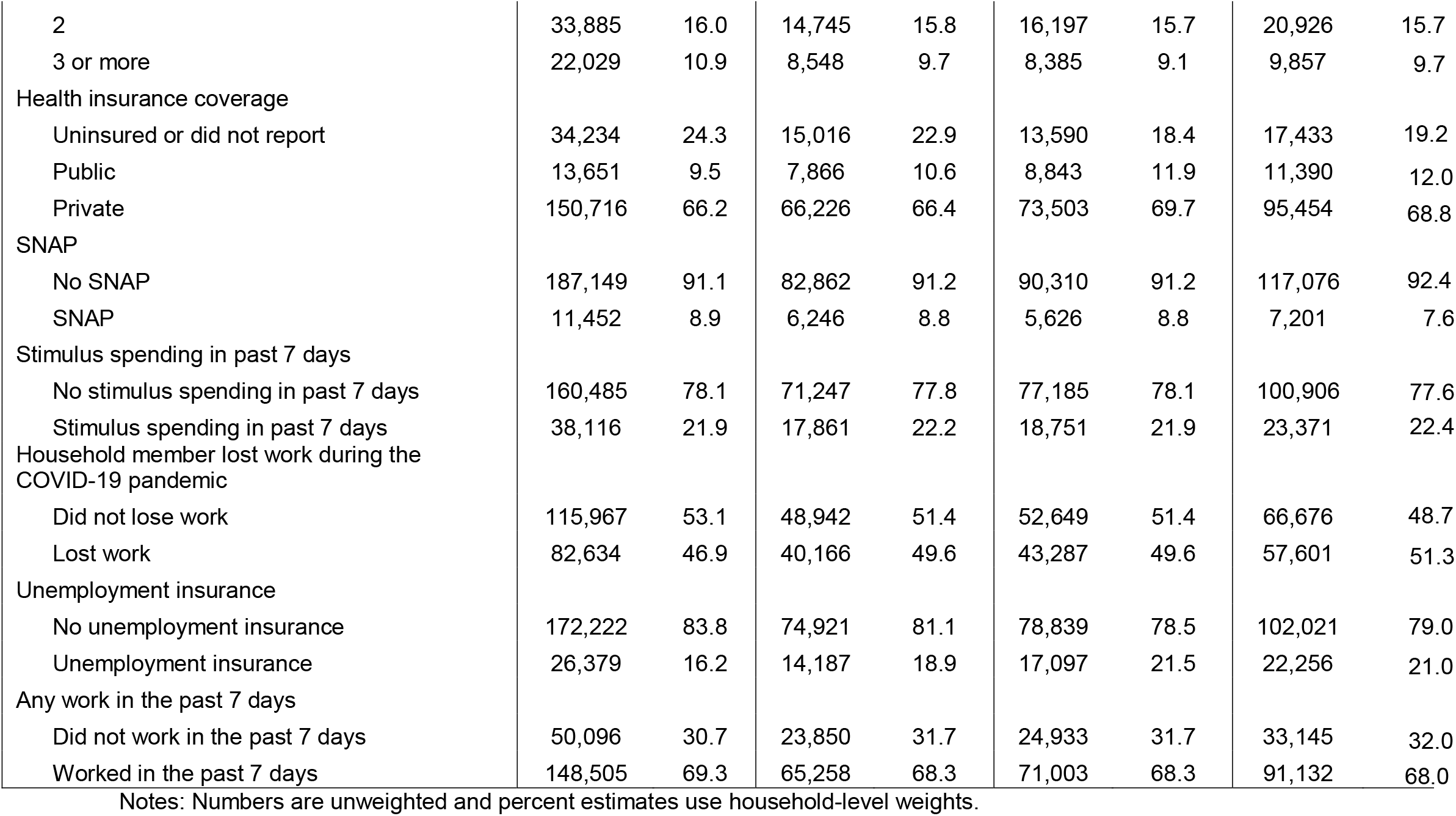
Participant characteristics in states with each minimum wage range.

In states with a minimum wage of less than $8.00, 14.3% of participants under age 65 and 16.6% of those in households with children reported household food insufficiency, while 10.3% of participants reported very low child food sufficiency (Figure 1). There were racial and ethnic disparities in household food insufficiency, with 24.3% of non-Hispanic Black participants’ households and 19.3% of Latinx participants’ households reporting household food insufficiency, relative to 11.0% of non-Hispanic White participants’ households. In states with a minimum wage of $12.00 or more, 11.8% of participants under age 65 and 13.8% of participants under age 65 in households with children reported household food insufficiency, while 8.9% of participants reported very low child food sufficiency. In states with a minimum wage of $12.00 or more, 18.2% of non-Hispanic Black, 17.9% of Latinx, and 8.6% of non-Hispanic White participants reported household food insufficiency.

**Figure 1:**
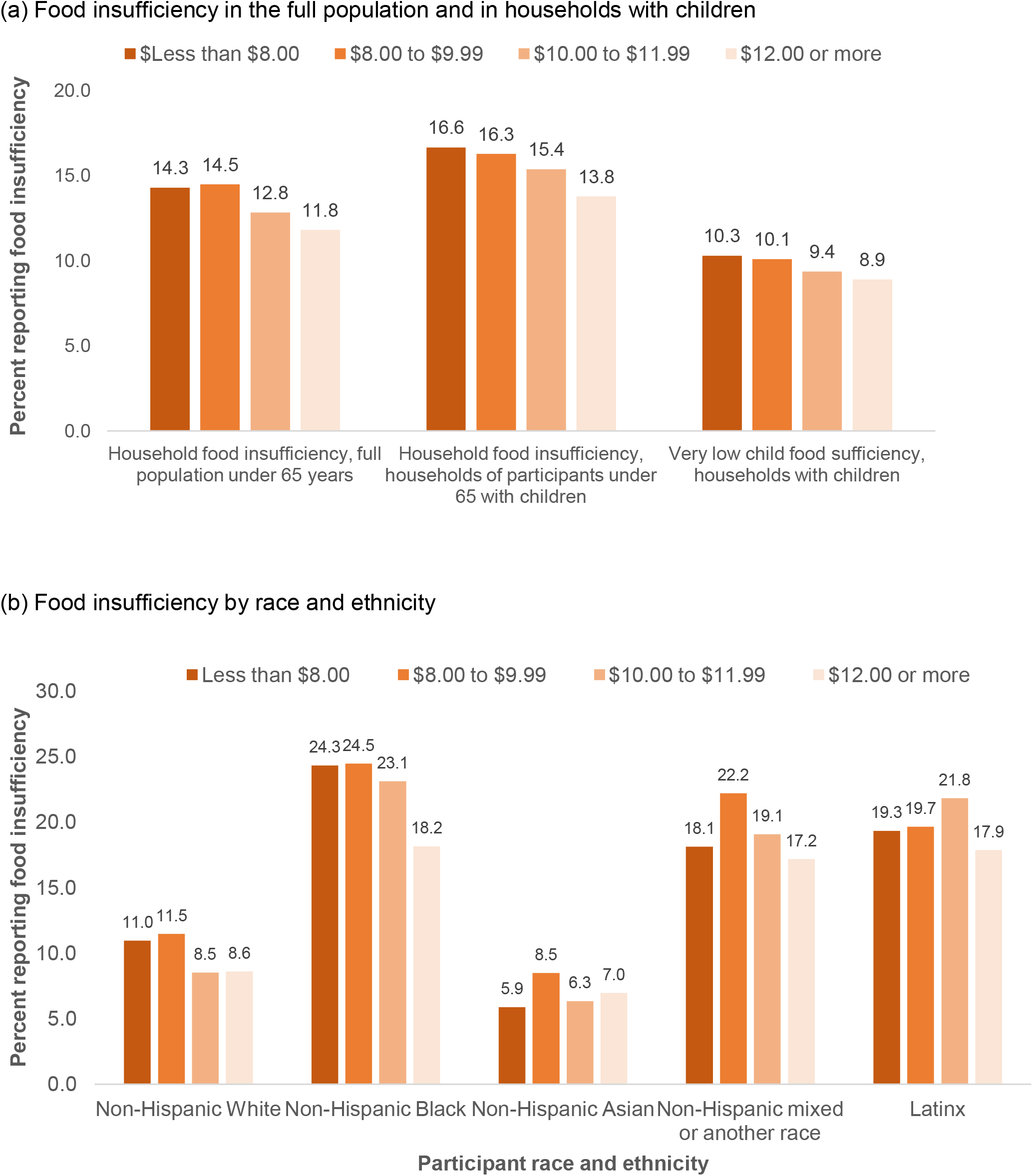
Percent of people under age 65 years reporting food insufficiency by state minimum wage. Note: Structural racism through policies such as slavery and redlining have shaped large racial disparities in wealth and in food insufficiency.

In the full sample of people under age 65, state minimum wage of $12 or more per hour was associated with a 1.83 percentage point reduction in the proportion of households reporting food insufficiency relative to a minimum wage of less than $8.00 per hour (95% CI: −2.67 to −0.99 percentage points, Table 2, Figure 2, Appendix Table 1). In households with children, a state minimum wage of $12 or more was associated with a 2.13 percentage point reduction in the proportion of households reporting food insufficiency (95% CI: −3.25 to −1.00 percentage points) and a 1.16 percentage point reduction in the proportion of households with children reporting very low child food sufficiency (95% CI: −1.69 to −0.63 percentage points). Minimum wages of $8.00 to $9.99 or $10.00 to $11.99 were not associated with changes in household food insufficiency overall or in households with children or with very low child food sufficiency.

**Table 2:**
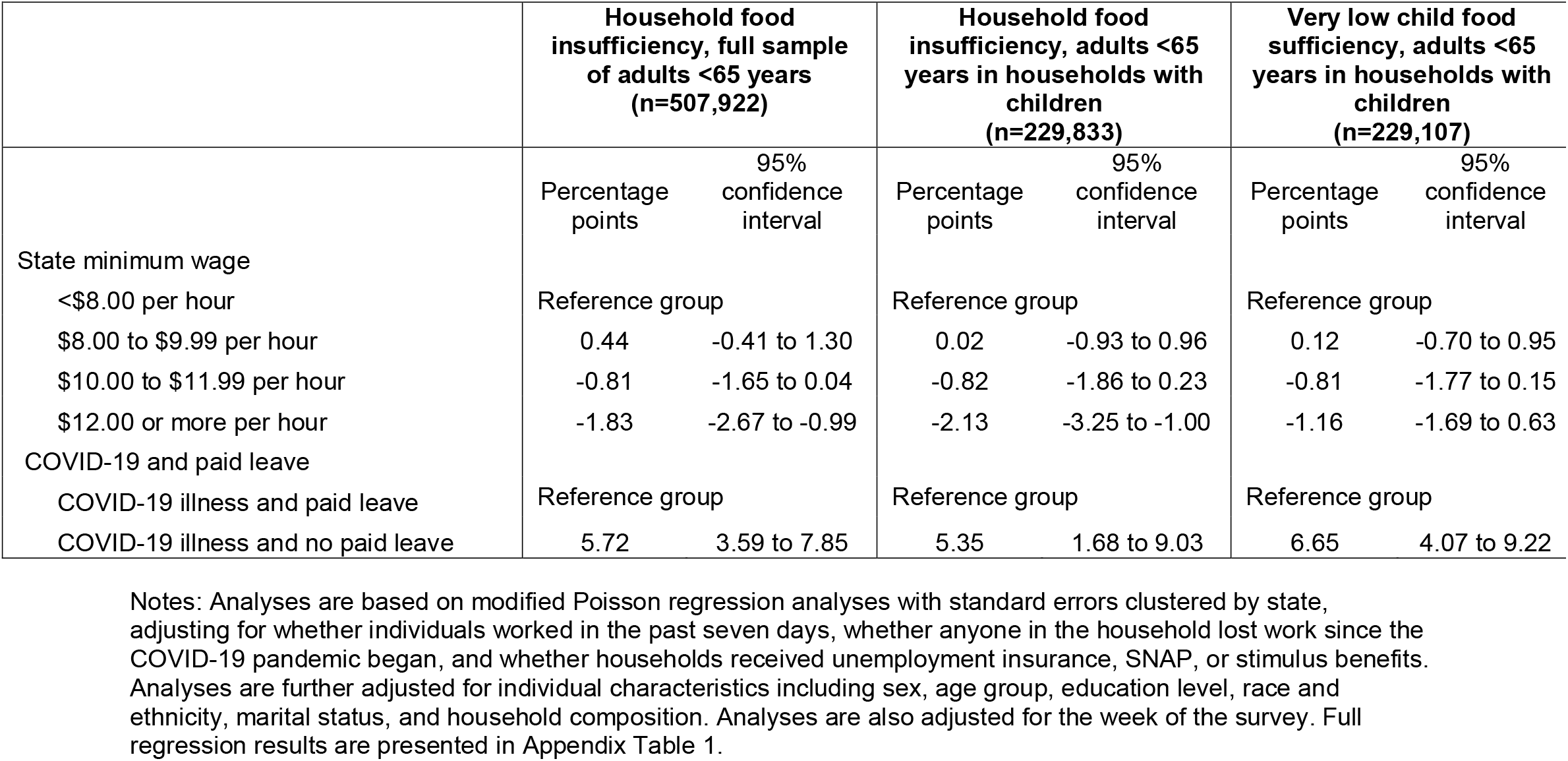
Poisson regression marginal effect estimates of the relationship between state minimum wage category and household food insufficiency.

|  | Household food insufficiency, full sample of adults <65 years (n=507,922) |  | Household food insufficiency, adults <65 years in households with children (n=229,833) |  | Very low child food sufficiency, adults <65 years in households with children (n=229,107) |  |
| --- | --- | --- | --- | --- | --- | --- |
|  | Percentage points | 95% confidence interval | Percentage points | 95% confidence interval | Percentage points | 95% confidence interval |
| State minimum wage |  |  |  |  |  |  |
| <\$8.00 per hour | Reference group | | Reference group | | Reference group | |
| \$8.00 to \$9.99 per hour | 0.44 | -0.41 to 1.30 | 0.02 | -0.93 to 0.96 | 0.12 | -0.70 to 0.95 |
| \$10.00 to \$11.99 per hour | -0.81 | -1.65 to 0.04 | -0.82 | -1.86 to 0.23 | -0.81 | -1.77 to 0.15 |
| \$12.00 or more per hour | -1.83 | -2.67 to -0.99 | -2.13 | -3.25 to -1.00 | -1.16 | -1.69 to 0.63 |
| COVID-19 and paid leave |  |  |  |  |  |  |
| COVID-19 illness and paid leave | Reference group |  | Reference group |  | Reference group |  |
| COVID-19 illness and no paid leave | 5.72 | 3.59 to 7.85 | 5.35 | 1.68 to 9.03 | 6.65 | 4.07 to 9.22 |

Notes: Analyses are based on modified Poisson regression analyses with standard errors clustered by state, adjusting for whether individuals worked in the past seven days, whether anyone in the household lost work since the COVID-19 pandemic began, and whether households received unemployment insurance, SNAP, or stimulus benefits. Analyses are further adjusted for individual characteristics including sex, age group, education level, race and ethnicity, marital status, and household composition. Analyses are also adjusted for the week of the survey. Full regression results are presented in Appendix Table 1.

**Figure 2:**
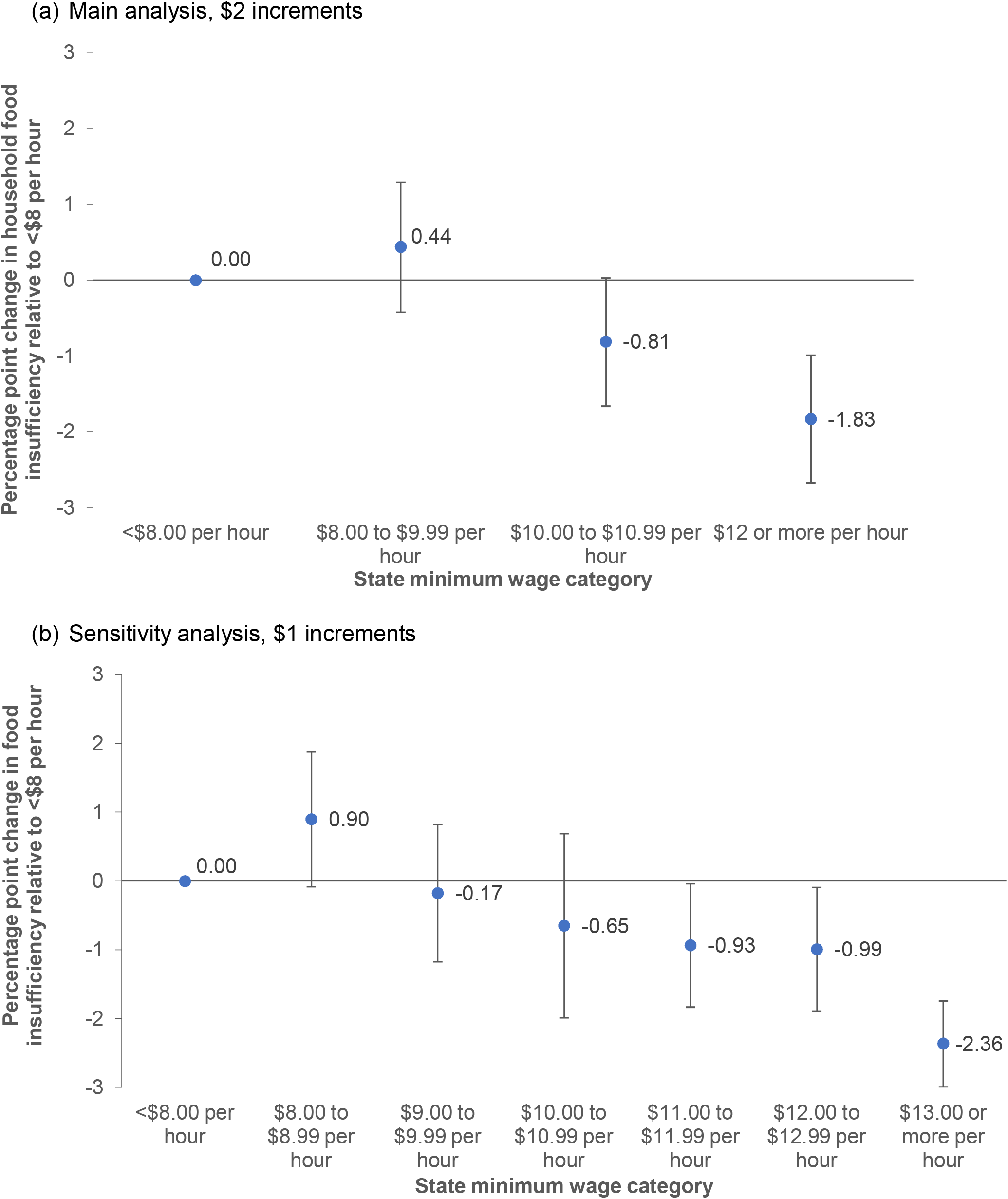
Marginal effect estimates of the relationship between state minimum wage, paid leave, and household food insufficiency among people younger than 65 years. Notes: Marginal effects estimates from multivariable modified Poisson regression analyses adjusting for all covariates (Appendix Tables 1 and 5).

Among 4,085 people who missed work due to COVID-19, 64.4% reported not receiving paid sick leave. Missing work due to COVID-19 was not associated with household food insufficiency in the full sample (0.47 percentage points, 95% CI: −0.98 to 1.92 percentage points) but was associated with a 5.72 percentage point increase in the proportion of households reporting food insufficiency for those who did not have paid leave (95% CI: 3.59 to 7.85 percentage points). In households with children, missing work due to COVID-19 without paid leave was associated with a 5.35 percentage point increase in the proportion of households reporting food insufficiency (95% CI: 1.68 to 9.03 percentage points) and a 6.65 percentage point increase in the proportion of households reporting very low child food sufficiency (95% CI: 4.07 to 9.22 percentage points).

In subgroup analyses by race and ethnicity (Table 3, Appendix Table 2), a state minimum wage of $12.00 or more per hour was associated with a reduction in the proportion of households with food insufficiency relative to a state minimum wage of $8.00 or less per hour among non-Hispanic Black people (−4.40 percentage points, 95% CI: −6.92 to −1.87 percentage points), and non-Hispanic White people (−1.83 percentage points, 95% CI: −2.40 to −1.26 percentage points). There was no relationship between state minimum wage and food insufficiency among Latinx people. State minimum wage of $8.00 to $9.99 or $10.00 to $11.99 was not associated with household food insufficiency relative to a state minimum wage of $8.00 or less per hour in these subgroups. Missing work due to COVID-19 infection without paid leave as associated with a 10.32 percentage point increase in the proportion of non-Hispanic Black participants reporting household food insufficiency (95% CI: 2.59 to 18.05 percentage points), a 9.55 percentage point increase in Latinx households reporting food insufficiency (95% CI: 3.02 to 16.06 percentage points), and a 4.47 percentage point increase in the proportion of non-Hispanic White households reporting food insufficiency (95% CI: 1.31 to 7.62 percentage points).

**Table 3:**
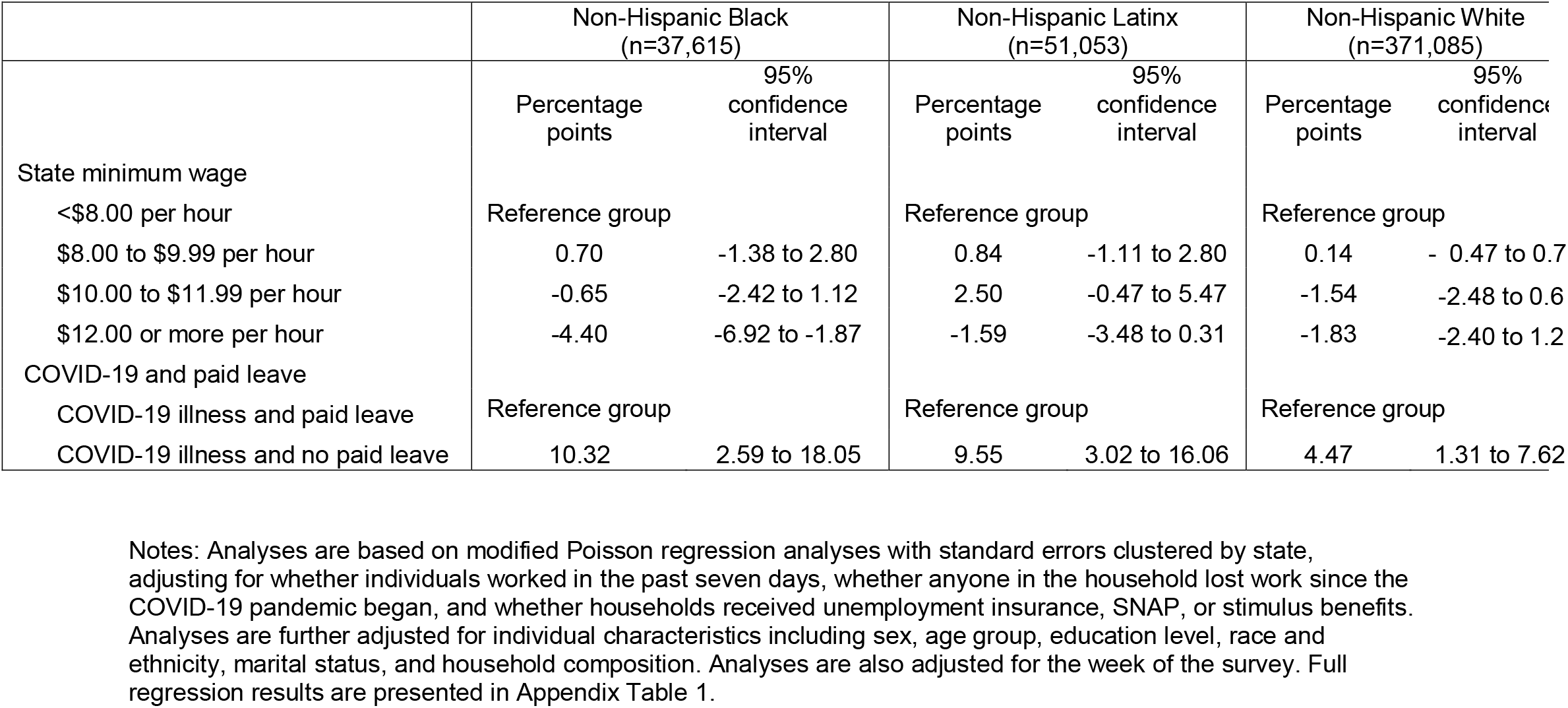
Poisson regression marginal effect estimates of the relationship between state minimum wage category and household food insufficiency, stratified by race/ethnicity.

Subgroup analyses among populations most likely to be affected by minimum wage policies were consistent with the main results (Appendix Table 3). Among participants who reported doing any work in the past seven days, a state minimum wage of $12.00 or more per hour was associated with a reduction in the proportion of households reporting food insufficiency (−1.61 percentage points, 95% CI: −2.16 to −1.06 percentage points) relative to a state minimum wage of $8.00 or less per hour. The results were consistent among participants living in households with a 2019 income of less than $75,000 (−2.08 percentage points, 95% CI: −3.17 to −0.98 percentage points) and among participants with a high school education or less (−2.54 percentage points, 95% CI: −4.12 to −0.95 percentage points). In all three subpopulations, state minimum wages of $8.00 to $9.99 or $10.00 to $11.99 were not associated with household food sufficiency.

We also conducted analyses of populations that should be less directly affected by minimum wage policies. Relative to a state minimum wage of less than $8, living in a state with a minimum wage of $12 or more was associated with smaller reductions in the proportions of households reporting food insufficiency among participants 65 years and older (−0.54 percentage points, 95% CI: −0.95 to −0.12 percentage points) and among those with a 2019 household income of $75,000 or more (−0.30 percentage points, 95% CI: −0.62 to 0.02 percentage points, Appendix Table 4).

Sensitivity analyses were consistent with the main result (Appendix Table 5) when we used $1 minimum wage increments as the main exposure variable, when we redefined food insufficiency as based on trouble paying for food, when we conducted the analysis without survey weights, and when we used balanced repeated replication to estimate standard errors rather than clustering standard errors by state.

## Discussion

In states with a minimum wage of less than $8.00, 14.1% of people under 65 years and 16.6% of households with children reported food insufficiency. Our results indicate that living in a state with at least a $12 minimum wage was associated with a reduction in the proportion of all households and households with children reporting food insufficiency during the COVID-19 pandemic among participants of working age. A $12 minimum wage was associated with a larger reduction in food insufficiency among non-Hispanic Black participants and may contribute to reducing racial disparities in food insufficiency. The main results were consistent with results in subgroups most likely to be directly affected by minimum wage policies, including those reporting working in the past seven days and with lower education and income. In populations less likely to be directly affected by state minimum wage policies − those over 65 years and of higher income - the association between state minimum wage policies and household food insufficiency was of a smaller magnitude.

We found that missing work due to COVID-19 without having paid sick leave was associated with increased food insufficiency, with larger effect estimates for households with children. The paid leave policies enacted by Congress in April 2020 excluded many workers and expired in 2020.^22^ Providing paid sick leave and safe food assistance for people who test positive for COVID-19 may prevent food insufficiency among people who are ill and their children, and could support people who become ill in staying home from work until they recover and complete quarantine. Prior evidence indicates that paid sick leave policies are associated with reduced onward transmission of infectious diseases.^47^

Our study has implications for further research. The relationship between state minimum wage and reduced food insufficiency was concentrated in states with a minimum wage of $12.00 or more. This suggests that evaluating state minimum wage policies that surpass this threshold rather than evaluating state minimum wages as a continuous variable may have relevance to future analyses of the relationship between state minimum wages and food insufficiency or other health and economic outcomes. There is also a need for further research on food insufficiency and economic precarity among tipped workers and independent contractors^48^ who may receive even less than the minimum wage.

### Strengths and Limitations

Strengths of our analyses include that the data are representative of all 50 US states. Additional strengths are that we conducted subgroup analyses in populations most likely to be directly affected by state minimum wage policies and falsification tests in populations less likely to be directly affected by state minimum wage policies. Our analyses were also consistent when redefined the exposure and outcome and took different approaches to weighting and to clustering standard errors. Finally, we were able to adjust for the receipt of several programs delivered by states that may be correlated with state minimum wage, including SNAP, health insurance, and unemployment insurance.

Our analysis was not causal, and it is likely that state minimum wage policies are correlated with other state-level social supports for people in low-income households. Given the short timeframe and the fact that only two states made changes to their minimum wages during the pandemic, we are unable to use state fixed effects to account for time-invariant state characteristics in this analysis. Further limitations of our study include that we did not have data on participants’ wages to determine which participants were receiving minimum wage and the low response rate to the Census Pulse survey. That there was an association, although of a smaller magnitude, between state minimum wage policies and food insufficiency in the falsification test among people over the age of 65 years could reflect unmeasured confounding by additional state policies that address food insufficiency, although it could also be due to some people over age 65 directly and indirectly affected by state minimum wage policies as workers or in the same household as workers.

## Conclusion

Food is essential for survival. The consequences of prolonged food insufficiency for a large proportion of the American population are likely to be severe. Improving food sufficiency for children in particular could improve their lifelong health and human capital.^10,49,50^ Our analyses indicates that there is a relationship between a minimum wage of $12.00 or more and reduced household food insufficiency overall and in households with children. We also found that missing work due to COVID-19 and not having paid sick leave was associated with increased food insufficiency, highlighting the importance of strengthening supports for paid leave. Policies increasing the minimum wage and providing paid sick leave for all workers may be associated with reduced food insufficiency for the working age population and for children, as well as reduce racial disparities in food insufficiency, as the United States begins to recover from the COVID-19 pandemic. Most of the US population supports increased minimum wage and paid leave policies.^51^

## Data Availability

Data are publicly available

https://tinyurl.com/statepolicies

https://datacatalog.urban.org/dataset/census-pulse-public-use-files-questionnaire-two

## Appendix

**Appendix Table 1:** Poisson regression estimates of the relationship between state minimum wage category and household food insufficiency.

|  | Household food insufficiency, full sample of adults <65 years (n=507,922) |  | Household food insufficiency, adults <65 years in households with children (n=229,833) |  | Very low child food sufficiency, adults <65 years in households with children (n=229,107) |  |
| --- | --- | --- | --- | --- | --- | --- |
|  | aPR | 95% confidence interval | aPR | 95% confidence interval | aPR | 95% confidence interval |
| State minimum wage |  |  |  |  |  |  |
| <\$8.00 per hour | Reference group | | Reference group | | Reference group | |
| \$8.00 to \$9.99 per hour | 1.032 | 0.972 - 1.096 | 1.001 | 0.945 - 1.060 | 1.012 | 0.934 - 1.097 |
| \$10.00 to \$11.99 per hour | 0.942 | 0.885 - 1.003 | 0.950 | 0.889 - 1.015 | 0.920 | 0.831 - 1.019 |
| \$12.00 or more per hour | 0.869 | 0.813 - 0.928 | 0.870 | 0.806 - 0.938 | 0.886 | 0.837 - 0.938 |
| Missed work due to COVID-19 and paid leave |  |  |  |  |  |  |
| COVID-19 illness and paid leave | Reference group |  | Reference group |  | Reference group |  |
| COVID-19 illness and no paid leave | 1.409 | 1.173 - 1.693 | 1.291 | 1.025 - 1.625 | 1.636 | 1.170 - 2.289 |
| Missed work due to COVID-19 |  |  |  |  |  |  |
| Did not miss work due to COVID-19 | Reference group |  | Reference group |  | Reference group |  |
| Missed work due to COVID-19 | 0.864 | 0.719 - 1.039 | 0.966 | 0.763 - 1.221 | 0.858 | 0.600 - 1.226 |
| Not working and paid sick leave |  |  |  |  |  |  |
| Not working and paid leave | Reference group |  | Reference group |  | Reference group |  |
| Not working and no paid leave | 1.071 | 0.998 - 1.149 | 1.057 | 0.932 - 1.199 | 1.128 | 0.990 - 1.285 |
| Race/ethnicity |  |  |  |  |  |  |
| Non-Hispanic White | Reference group |  | Reference group |  | Reference group |  |
| Non-Hispanic Black | 1.431 | 1.361 - 1.504 | 1.433 | 1.342 - 1.530 | 1.791 | 1.589 - 2.019 |
| Latinx | 1.209 | 1.124 - 1.301 | 1.191 | 1.123 - 1.262 | 1.468 | 1.345 - 1.601 |
| Non-Hispanic Asian | 0.885 | 0.739 - 1.060 | 0.921 | 0.718 - 1.180 | 1.243 | 0.930 - 1.662 |
| Non-Hispanic mixed race or another race | 1.351 | 1.264 - 1.444 | 1.388 | 1.289 - 1.494 | 1.603 | 1.419 - 1.810 |
| Gender |  |  |  |  |  |  |
| Man | Reference group |  | Reference group |  | Reference group |  |
| Woman | 1.056 | 1.030 - 1.082 | 1.071 | 1.005 - 1.140 | 1.098 | 1.009 - 1.194 |
| Age group |  |  |  |  |  |  |
| 18 to 24 | Reference group |  | Reference group |  | Reference group |  |
| 25 to 39 | 1.288 | 1.196 - 1.387 | 1.628 | 1.397 - 1.897 | 1.699 | 1.363 - 2.118 |
| 40 to 54 | 1.336 | 1.243 - 1.435 | 1.620 | 1.399 - 1.875 | 1.856 | 1.470 - 2.344 |
| 55 to 64 | 0.977 | 0.884 - 1.080 | 1.388 | 1.173 - 1.643 | 1.484 | 1.173 - 1.877 |
| Education |  |  |  |  |  |  |
| < High school graduate | Reference group |  | Reference group |  | Reference group |  |
| High school grad | 0.833 | 0.785 - 0.884 | 0.888 | 0.818 - 0.964 | 0.913 | 0.840 - 0.992 |
| Some college | 0.646 | 0.614 - 0.679 | 0.695 | 0.643 - 0.751 | 0.712 | 0.648 - 0.782 |
| College graduate or greater | 0.281 | 0.262 - 0.301 | 0.293 | 0.267 - 0.322 | 0.315 | 0.276 - 0.359 |
| Marital status |  |  |  |  |  |  |
| Unmarried | Reference group |  | Reference group |  | Reference group |  |
| Married | 0.684 | 0.656 - 0.713 | 0.688 | 0.651 - 0.727 | 0.686 | 0.633 - 0.744 |
| Number of adults in household |  |  |  |  |  |  |
| 1 | Reference group |  | Reference group |  | Reference group |  |
| 2 | 0.811 | 0.781 - 0.841 | 0.829 | 0.785 - 0.874 | 0.794 | 0.734 - 0.859 |
| 3 | 0.798 | 0.766 - 0.832 | 0.908 | 0.855 - 0.963 | 0.894 | 0.822 - 0.971 |
| 4 or more | 0.854 | 0.825 - 0.884 | 0.969 | 0.896 - 1.048 | 0.903 | 0.806 - 1.011 |
| Number of children in household |  |  |  |  |  |  |
| 0 | Reference group |  | N/A |  | N/A |  |
| 1 | 1.064 | 1.014 - 1.116 | Reference group |  | Reference group |  |
| 2 | 1.047 | 1.001 - 1.094 | 0.995 | 0.934 - 1.060 | 1.120 | 1.049 - 1.196 |
| 3 or more | 1.167 | 1.110 - 1.227 | 1.139 | 1.082 - 1.199 | 1.377 | 1.284 - 1.476 |
| Health insurance coverage |  |  |  |  |  |  |
| Uninsured or did not report | Reference group |  | Reference group |  | Reference group |  |
| Public | 1.025 | 0.983 - 1.067 | 1.031 | 0.952 - 1.117 | 1.073 | 0.986 - 1.168 |
| Private | 0.652 | 0.632 - 0.672 | 0.723 | 0.692 - 0.755 | 0.779 | 0.730 - 0.832 |
| SNAP |  |  |  |  |  |  |
| No SNAP | Reference group |  | Reference group |  | Reference group |  |
| SNAP | 1.250 | 1.186 - 1.316 | 1.149 | 1.080 - 1.221 | 1.129 | 1.033 - 1.235 |
| Stimulus spending in past 7 days |  |  |  |  |  |  |
| No stimulus spending in past 7 days | Reference group |  | Reference group |  | Reference group |  |
| Stimulus spending in past 7 days | 1.212 | 1.170 - 1.255 | 1.190 | 1.127 - 1.256 | 1.214 | 1.122 - 1.312 |
| Household member lost work during the COVID-19 pandemic |  |  |  |  |  |  |
| Did not lose work | Reference group |  | Reference group |  | Reference group |  |
| Lost work | 2.204 | 2.077 - 2.339 | 2.095 | 1.953 - 2.247 | 2.384 | 2.214 - 2.568 |
| Unemployment insurance |  |  |  |  |  |  |
| No unemployment insurance | Reference group |  | Reference group |  | Reference group |  |
| Unemployment insurance | 0.910 | 0.873 - 0.949 | 0.926 | 0.881 - 0.974 | 0.891 | 0.824 - 0.963 |
| Any work in the past 7 days |  |  |  |  |  |  |
| Did not work in the past 7 days | Reference group |  | Reference group |  | Reference group |  |
| Worked in the past 7 days | 0.697 | 0.639 - 0.760 | 0.739 | 0.642 - 0.851 | 0.765 | 0.652 - 0.897 |
| Wave and date |  |  |  |  |  |  |
| Wave 13, 8/19 to 8/31 | Reference group |  | Reference group |  | Reference group |  |
| Wave 14, 9/2 to 9/14 | 1.003 | 0.934 - 1.076 | 1.010 | 0.934 - 1.093 | 0.967 | 0.866 - 1.078 |
| Wave 15, 9/16 to 9/28 | 0.979 | 0.911 - 1.051 | 0.977 | 0.877 - 1.088 | 0.957 | 0.820 - 1.117 |
| Wave 16, 9/30 to 10/12 | 1.002 | 0.941 - 1.067 | 1.045 | 0.959 - 1.138 | 1.043 | 0.947 - 1.149 |
| Wave 17, 10/14 to 10/26 | 1.057 | 0.970 - 1.151 | 1.052 | 0.943 - 1.174 | 1.000 | 0.861 - 1.162 |
| Wave 18, 10/28 to 11/9 | 1.129 | 1.016 - 1.253 | 1.075 | 0.948 - 1.220 | 1.020 | 0.873 - 1.192 |
| Wave 19, 11/11 to 11/23 | 1.107 | 1.005 - 1.219 | 1.110 | 1.002 - 1.230 | 1.083 | 0.950 - 1.234 |
| Wave 20, 11/25 to 12/7 | 1.174 | 1.084 - 1.270 | 1.176 | 1.048 - 1.319 | 1.217 | 1.068 - 1.386 |
| Wave 21, 12/9 to 12/21 | 1.231 | 1.122 - 1.351 | 1.239 | 1.105 - 1.390 | 1.226 | 1.085 - 1.385 |

**Appendix Table 2:** Poisson regression estimates of the relationship between state minimum wage category and household food insufficiency, stratified by race/ethnicity.

|  | Non-Hispanic Black<br>(n=37,615) |  | Non-Hispanic Latinx<br>(n=51,053) |  | Non-Hispanic White<br>(n=371,085) |  |
| --- | --- | --- | --- | --- | --- | --- |
|  | aPR | 95%<br>confidence<br>interval | aPR | 95%<br>confidence<br>interval | aPR | 95%<br>confidence<br>interval |
| State minimum wage |  |  |  |  |  |  |
| <\$8.00 per hour | Reference group | | Reference group | | Reference group | |
| \$8.00 to \$9.99 per hour | 1.030 | 0.945 - 1.122 | 1.044 | 0.946 - 1.152 | 1.013 | 0.957 - 1.071 |
| \$10.00 to \$11.99 per hour | 0.973 | 0.902 - 1.049 | 1.130 | 0.983 - 1.299 | 0.857 | 0.775 - 0.947 |
| \$12.00 or more per hour | 0.816 | 0.721 - 0.924 | 0.918 | 0.827 - 1.019 | 0.829 | 0.782 - 0.880 |
| Missed work due to COVID-19 and paid leave |  |  |  |  |  |  |
| COVID-19 illness and paid leave |  |  |  |  |  |  |
| COVID-19 illness and no paid leave | 1.576 | 0.998 - 2.489 | 1.397 | 1.082 - 1.805 | 1.426 | 1.003 - 2.027 |
| Missed work due to COVID-19 |  |  |  |  |  |  |
| Did not miss work due to COVID-19 |  |  |  |  |  |  |
| Missed work due to COVID-19 | 0.748 | 0.453 - 1.235 | 0.892 | 0.753 - 1.055 | 0.862 | 0.630 - 1.180 |
| Not working and paid sick leave |  |  |  |  |  |  |
| Not working and paid leave |  |  |  |  |  |  |
| Not working and no paid leave | 1.009 | 0.888 - 1.148 | 1.142 | 1.028 - 1.268 | 1.072 | 0.960 - 1.196 |
| Gender |  |  |  |  |  |  |
| Man | Reference group |  | Reference group |  | Reference group |  |
| Woman | 1.032 | 0.951 - 1.120 | 0.968 | 0.888 - 1.054 | 1.130 | 1.087 - 1.175 |
| Age group |  |  |  |  |  |  |
| 18 to 24 | Reference group |  | Reference group |  | Reference group |  |
| 25 to 39 | 1.237 | 1.033 - 1.480 | 1.340 | 1.081 - 1.661 | 1.226 | 1.125 - 1.337 |
| 40 to 54 | 1.167 | 0.967 - 1.408 | 1.467 | 1.200 - 1.794 | 1.259 | 1.132 - 1.401 |
| 55 to 64 | 0.826 | 0.687 - 0.994 | 1.272 | 1.103 - 1.467 | 0.855 | 0.764 - 0.957 |
| Education |  |  |  |  |  |  |
| < High school graduate | Reference group |  | Reference group |  | Reference group |  |
| High school grad | 1.013 | 0.882 - 1.164 | 0.836 | 0.755 - 0.925 | 0.729 | 0.669 - 0.794 |
| Some college | 0.830 | 0.723 - 0.953 | 0.658 | 0.595 - 0.727 | 0.551 | 0.509 - 0.596 |
| College graduate or greater | 0.441 | 0.384 - 0.506 | 0.350 | 0.300 - 0.409 | 0.213 | 0.193 - 0.235 |
| Marital status |  |  |  |  |  |  |
| Unmarried | Reference group |  | Reference group |  | Reference group |  |
| Married | 0.764 | 0.717 - 0.814 | 0.758 | 0.689 - 0.834 | 0.633 | 0.607 - 0.661 |
| Number of adults in household |  |  |  |  |  |  |
| 1 | Reference group |  | Reference group |  | Reference group |  |
| 2 | 0.845 | 0.777 - 0.919 | 0.759 | 0.682 - 0.846 | 0.816 | 0.786 - 0.847 |
| 3 | 0.801 | 0.727 - 0.883 | 0.752 | 0.691 - 0.817 | 0.796 | 0.755 - 0.840 |
| 4 or more | 0.848 | 0.753 - 0.957 | 0.768 | 0.671 - 0.880 | 0.872 | 0.814 - 0.935 |
| Number of children in household |  |  |  |  |  |  |
| 0 | Reference group |  | Reference group |  | Reference group |  |
| 1 | 1.151 | 1.073 - 1.234 | 1.034 | 0.974 - 1.098 | 1.024 | 0.971 - 1.080 |
| 2 | 1.061 | 0.982 - 1.146 | 1.043 | 0.939 - 1.159 | 1.048 | 1.007 - 1.089 |
| 3 or more | 1.166 | 1.032 - 1.317 | 1.181 | 1.100 - 1.267 | 1.175 | 1.094 - 1.263 |
| Health insurance coverage |  |  |  |  |  |  |
| Uninsured or did not report | Reference group |  | Reference group |  | Reference group |  |
| Public | 0.935 | 0.866 - 1.010 | 0.996 | 0.938 - 1.058 | 1.089 | 1.031 - 1.150 |
| Private | 0.783 | 0.707 - 0.867 | 0.726 | 0.689 - 0.765 | 0.591 | 0.569 - 0.615 |
| SNAP |  |  |  |  |  |  |
| No SNAP | Reference group |  | Reference group |  | Reference group |  |
| SNAP | 0.997 | 0.927 - 1.072 | 1.283 | 1.159 - 1.419 | 1.418 | 1.325 - 1.518 |
| Stimulus spending in past 7 days |  |  |  |  |  |  |
| No stimulus spending in past 7 days | Reference group |  | Reference group |  | Reference group |  |
| Stimulus spending in past 7 days | 1.146 | 1.076 - 1.221 | 1.053 | 1.018 - 1.090 | 1.317 | 1.269 - 1.366 |
| Household member lost work during the COVID-19 pandemic |  |  |  |  |  |  |
| Did not lose work | Reference group |  | Reference group |  | Reference group |  |
| Lost work | 1.824 | 1.689 - 1.969 | 2.011 | 1.782 - 2.270 | 2.488 | 2.299 - 2.691 |
| Unemployment insurance |  |  |  |  |  |  |
| No unemployment insurance | Reference group |  | Reference group |  | Reference group |  |
| Unemployment insurance | 0.897 | 0.848 - 0.949 | 0.859 | 0.800 - 0.922 | 0.936 | 0.884 - 0.990 |
| Any work in the past 7 days |  |  |  |  |  |  |
| Did not work in the past 7 days | Reference group |  | Reference group |  | Reference group |  |
| Worked in the past 7 days | 0.701 | 0.601 - 0.818 | 0.761 | 0.659 - 0.878 | 0.687 | 0.616 - 0.765 |
| Wave and date |  |  |  |  |  |  |
| Wave 13, 8/19 to 8/31 | Reference group |  | Reference group |  | Reference group |  |
| Wave 14, 9/2 to 9/14 | 0.974 | 0.892 - 1.064 | 1.012 | 0.940 - 1.091 | 1.018 | 0.910 - 1.139 |
| Wave 15, 9/16 to 9/28 | 0.899 | 0.809 - 0.999 | 0.992 | 0.832 - 1.183 | 1.018 | 0.934 - 1.109 |
| Wave 16, 9/30 to 10/12 | 0.987 | 0.887 - 1.099 | 0.939 | 0.862 - 1.023 | 1.046 | 0.957 - 1.142 |
| Wave 17, 10/14 to 10/26 | 1.059 | 0.956 - 1.173 | 0.999 | 0.850 - 1.174 | 1.100 | 0.987 - 1.227 |
| Wave 18, 10/28 to 11/9 | 1.072 | 0.906 - 1.267 | 1.165 | 0.999 - 1.359 | 1.181 | 1.064 - 1.311 |
| Wave 19, 11/11 to 11/23 | 1.084 | 0.965 - 1.217 | 0.990 | 0.806 - 1.218 | 1.180 | 1.065 - 1.307 |
| Wave 20, 11/25 to 12/7 | 1.133 | 0.971 - 1.322 | 1.164 | 0.998 - 1.359 | 1.230 | 1.111 - 1.363 |
| Wave 21, 12/9 to 12/21 | 1.226 | 1.079 - 1.393 | 1.125 | 0.992 - 1.276 | 1.325 | 1.193 - 1.471 |

**Appendix Table 3:** Analyses of subgroup most directly affected by state minimum wage policies and food insufficiency.

| | Reported any work in the past week<br>(n=375,898) | | 2019 household income <\$75,000<br>(n=264,671) | | Education of high school graduate or less<br>(n=66,601) | |
| --- | --- | --- | --- | --- | --- | --- |
|  | aPR | 95% confidence interval | aPR | 95% confidence interval | aPR | 95% confidence interval |
| State minimum wage |  |  |  |  |  |  |
| <\$8.00 per hour | Reference group | | Reference group | | Reference group | |
| \$8.00 to \$9.99 per hour | 1.022 | 0.950 - 1.099 | 1.017 | 0.960 - 1.076 | 1.032 | 0.947 - 1.125 |
| \$10.00 to \$11.99 per hour | 0.947 | 0.873 - 1.027 | 0.959 | 0.910 - 1.012 | 0.970 | 0.901 - 1.044 |
| \$12.00 or more per hour | 0.827 | 0.775 - 0.883 | 0.896 | 0.844 - 0.950 | 0.887 | 0.822 - 0.957 |
| Missed work due to COVID-19 and paid leave |  |  |  |  |  |  |
| COVID-19 illness and paid leave | N/A |  | Reference group |  | Reference group |  |
| COVID-19 illness and no paid leave |  |  | 0.817 | 0.660 - 1.012 | 0.871 | 0.665 - 1.141 |
| Missed work due to COVID-19 |  |  |  |  |  |  |
| Did not miss work due to COVID-19 | N/A |  | Reference group |  | Reference group |  |
| Missed work due to COVID-19 |  |  | 1.085 | 1.020 - 1.155 | 1.080 | 0.971 - 1.200 |
| Not working and paid sick leave |  |  |  |  |  |  |
| Not working and paid leave | N/A |  | Reference group |  | Reference group |  |
| Not working and no paid leave |  |  | 1.497 | 1.206 - 1.857 | 1.426 | 1.109 - 1.833 |
| Race/ethnicity |  |  |  |  |  |  |
| Non-Hispanic White | Reference group |  | Reference group |  | Reference group |  |
| Non-Hispanic Black | 1.640 | 1.544 - 1.742 | 1.332 | 1.264 - 1.403 | 1.286 | 1.208 - 1.370 |
| Latinx | 1.265 | 1.129 - 1.418 | 1.133 | 1.052 - 1.220 | 1.131 | 1.025 - 1.248 |
| Non-Hispanic Asian | 0.871 | 0.706 - 1.075 | 0.848 | 0.714 - 1.006 | 0.937 | 0.727 - 1.207 |
| Non-Hispanic mixed race or another race | 1.319 | 1.189 - 1.464 | 1.289 | 1.209 - 1.375 | 1.203 | 1.105 - 1.309 |
| Gender |  |  |  |  |  |  |
| Man | Reference group |  | Reference group |  | Reference group |  |
| Woman | 1.182 | 1.127 - 1.240 | 1.025 | 0.997 - 1.054 | 1.062 | 1.026 - 1.099 |
| Age group |  |  |  |  |  |  |
| 18 to 24 | Reference group |  | Reference group |  | Reference group |  |
| 25 to 39 | 1.103 | 1.007 - 1.208 | 1.288 | 1.179 - 1.407 | 1.226 | 1.090 - 1.378 |
| 40 to 54 | 1.068 | 0.956 - 1.194 | 1.368 | 1.262 - 1.484 | 1.232 | 1.120 - 1.355 |
| 55 to 64 | 0.711 | 0.608 - 0.831 | 1.002 | 0.901 - 1.114 | 0.856 | 0.757 - 0.969 |
| Education |  |  |  |  |  |  |
| < High school graduate | Reference group |  | Reference group |  | Reference group |  |
| High school grad | 0.808 | 0.692 - 0.942 | 0.844 | 0.790 - 0.901 | 0.807 | 0.759 - 0.858 |
| Some college | 0.600 | 0.528 - 0.682 | 0.679 | 0.642 - 0.719 | N/A |  |
| College graduate or greater | 0.221 | 0.193 - 0.252 | 0.366 | 0.339 - 0.395 | N/A |  |
| Marital status |  |  |  |  |  |  |
| Unmarried | Reference group |  | Reference group |  | Reference group |  |
| Married | 0.629 | 0.593 - 0.667 | 0.763 | 0.736 - 0.792 | 0.729 | 0.691 - 0.769 |
| Number of adults in household |  |  |  |  |  |  |
| 1 | Reference group |  | Reference group |  | Reference group |  |
| 2 | 0.765 | 0.709 - 0.825 | 0.865 | 0.835 - 0.896 | 0.837 | 0.789 - 0.888 |
| 3 | 0.742 | 0.688 - 0.801 | 0.832 | 0.795 - 0.870 | 0.822 | 0.770 - 0.878 |
| 4 or more | 0.811 | 0.751 - 0.876 | 0.891 | 0.855 - 0.928 | 0.877 | 0.831 - 0.925 |
| Number of children in household |  |  |  |  |  |  |
| 0 | Reference group |  | Reference group |  | Reference group |  |
| 1 | 1.115 | 1.033 - 1.203 | 1.071 | 1.023 - 1.121 | 1.026 | 0.964 - 1.092 |
| 2 | 1.161 | 1.080 - 1.248 | 1.037 | 0.983 - 1.094 | 1.018 | 0.937 - 1.105 |
| 3 or more | 1.321 | 1.232 - 1.415 | 1.144 | 1.082 - 1.210 | 1.108 | 1.031 - 1.191 |
| Health insurance coverage |  |  |  |  |  |  |
| Uninsured or did not report | Reference group |  | Reference group |  | Reference group |  |
| Public | 1.041 | 0.955 - 1.136 | 1.060 | 1.019 - 1.103 | 0.999 | 0.934 - 1.069 |
| Private | 0.671 | 0.641 - 0.702 | 0.792 | 0.767 - 0.817 | 0.688 | 0.649 - 0.728 |
| SNAP |  |  |  |  |  |  |
| No SNAP | Reference group |  | Reference group |  | Reference group |  |
| SNAP | 1.285 | 1.166 - 1.416 | 1.209 | 1.147 - 1.274 | 1.191 | 1.126 - 1.260 |
| Stimulus spending in past 7 days |  |  |  |  |  |  |
| No stimulus spending in past 7 days | Reference group |  | Reference group |  | Reference group |  |
| Stimulus spending in past 7 days | 1.368 | 1.298 - 1.441 | 1.138 | 1.096 - 1.182 | 1.133 | 1.064 - 1.208 |
| Household member lost work during the COVID-19 pandemic |  |  |  |  |  |  |
| Did not lose work | Reference group |  | Reference group |  | Reference group |  |
| Lost work | 2.413 | 2.288 - 2.545 | 2.045 | 1.936 - 2.162 | 1.897 | 1.755 - 2.051 |
| Unemployment insurance |  |  |  |  |  |  |
| No unemployment insurance | Reference group |  | Reference group |  | Reference group |  |
| Unemployment insurance | 0.898 | 0.858 - 0.939 | 0.894 | 0.855 - 0.934 | 0.890 | 0.839 - 0.945 |
| Any work in the past 7 days |  |  |  |  |  |  |
| Did not work in the past 7 days | Reference group |  | Reference group |  | Reference group |  |
| Worked in the past 7 days | N/A |  | 0.740 | 0.687 - 0.796 | 0.723 | 0.636 - 0.822 |
| Wave and date |  |  |  |  |  |  |
| Wave 13, 8/19 to 8/31 | Reference group |  | Reference group |  | Reference group |  |
| Wave 14, 9/2 to 9/14 | 0.989 | 0.898 - 1.089 | 1.007 | 0.943 - 1.074 | 0.943 | 0.855 - 1.039 |
| Wave 15, 9/16 to 9/28 | 0.980 | 0.898 - 1.068 | 0.976 | 0.906 - 1.051 | 0.939 | 0.848 - 1.040 |
| Wave 16, 9/30 to 10/12 | 0.929 | 0.845 - 1.022 | 0.994 | 0.930 - 1.062 | 0.985 | 0.902 - 1.076 |
| Wave 17, 10/14 to 10/26 | 1.016 | 0.895 - 1.154 | 1.052 | 0.962 - 1.151 | 1.032 | 0.923 - 1.153 |
| Wave 18, 10/28 to 11/9 | 1.165 | 1.026 - 1.322 | 1.135 | 1.021 - 1.262 | 1.092 | 0.942 - 1.265 |
| Wave 19, 11/11 to 11/23 | 1.091 | 0.947 - 1.257 | 1.085 | 0.975 - 1.206 | 1.049 | 0.914 - 1.205 |
| Wave 20, 11/25 to 12/7 | 1.164 | 1.072 - 1.264 | 1.159 | 1.078 - 1.247 | 1.109 | 0.994 - 1.236 |
| Wave 21, 12/9 to 12/21 | 1.306 | 1.166 - 1.461 | 1.224 | 1.118 - 1.339 | 1.186 | 1.050 - 1.340 |

**Appendix Table 4:** Falsification tests, state minimum wage policies and food insufficiency.

| | **65 years or older & ≤2 adults in household**<br>**(n=186,424)** | | **2019 household income of \$75,000 or more**<br>**(n=243,251)** | |
| --- | --- | --- | --- | --- |
|  | aPR | 95% confidence interval | aPR | 95% confidence interval |
| State minimum wage |  |  |  |  |
| <\$8.00 per hour | Reference group | | Reference group | |
| \$8.00 to \$9.99 per hour | 1.102 | 0.966 - 1.257 | 1.162 | 0.988 - 1.367 |
| \$10.00 to \$11.99 per hour | 0.914 | 0.828 - 1.008 | 0.996 | 0.840 - 1.181 |
| \$12.00 or more per hour | 0.880 | 0.797 - 0.970 | 0.903 | 0.809 - 1.008 |
| Missed work due to COVID-19 and paid leave |  |  |  |  |
| COVID-19 illness and paid leave | Reference group |  | Reference group |  |
| COVID-19 illness and no paid leave | 0.910 | 0.382 - 2.169 | 0.887 | 0.411 - 1.916 |
| Missed work due to COVID-19 |  |  |  |  |
| Did not miss work due to COVID-19 | Reference group |  | Reference group |  |
| Missed work due to COVID-19 | 1.869 | 0.878 - 3.980 | 1.279 | 0.611 - 2.676 |
| Not working and paid sick leave |  |  |  |  |
| Not working and paid leave | Reference group |  | Reference group |  |
| Not working and no paid leave | 1.224 | 0.931 - 1.607 | 1.087 | 0.779 - 1.518 |
| Race/ethnicity |  |  |  |  |
| Non-Hispanic White | Reference group |  | Reference group |  |
| Non-Hispanic Black | 1.550 | 1.342 - 1.790 | 1.865 | 1.616 - 2.153 |
| Latinx | 1.827 | 1.616 - 2.067 | 1.261 | 1.004 - 1.584 |
| Non-Hispanic Asian | 1.129 | 0.889 - 1.434 | 1.013 | 0.766 - 1.341 |
| Non-Hispanic mixed race or another race | 1.530 | 1.212 - 1.930 | 1.562 | 1.287 - 1.897 |
| Gender |  |  |  |  |
| Man | Reference group |  | Reference group |  |
| Woman | 0.834 | 0.735 - 0.946 | 0.987 | 0.918 - 1.062 |
| Age group |  |  |  |  |
| 18 to 24 | N/A |  | Reference group |  |
| 25 to 39 |  |  | 1.841 | 1.226 - 2.765 |
| 40 to 54 |  |  | 1.980 | 1.290 - 3.037 |
| 55 to 64 |  |  | 1.311 | 0.822 - 2.090 |
| Education |  |  |  |  |
| < High school graduate | Reference group |  | Reference group |  |
| High school grad | 0.639 | 0.524 - 0.779 | 0.772 | 0.602 - 0.990 |
| Some college | 0.562 | 0.458 - 0.689 | 0.628 | 0.467 - 0.843 |
| College graduate or greater | 0.318 | 0.259 - 0.389 | 0.297 | 0.227 - 0.389 |
| Marital status |  |  |  |  |
| Unmarried | Reference group |  | Reference group |  |
| Married | 0.457 | 0.414 - 0.504 | 0.638 | 0.546 - 0.746 |
| Number of adults in household |  |  |  |  |
| 1 | Reference group |  | Reference group |  |
| 2 | 0.838 | 0.730 - 0.962 | 0.667 | 0.564 - 0.789 |
| 3 | 1.054 | 0.854 - 1.302 | 0.822 | 0.691 - 0.977 |
| 4 or more | 1.199 | 0.963 - 1.493 | 0.941 | 0.784 - 1.130 |
| Number of children in household |  |  |  |  |
| 0 | Reference group |  | Reference group |  |
| 1 | 1.307 | 1.104 - 1.548 | 1.120 | 0.948 - 1.324 |
| 2 | 1.398 | 1.189 - 1.644 | 1.448 | 1.286 - 1.631 |
| 3 or more | 1.467 | 1.038 - 2.075 | 1.737 | 1.533 - 1.969 |
| Health insurance coverage |  |  |  |  |
| Uninsured or did not report | Reference group |  | Reference group |  |
| Public | 1.005 | 0.896 - 1.127 | 0.796 | 0.657 - 0.964 |
| Private | 0.634 | 0.565 - 0.710 | 0.496 | 0.411 - 0.600 |
| SNAP |  |  |  |  |
| No SNAP | Reference group |  | Reference group |  |
| SNAP | 1.853 | 1.589 - 2.160 | 1.950 | 1.619 - 2.349 |
| Stimulus spending in past 7 days |  |  |  |  |
| No stimulus spending in past 7 days | Reference group |  | Reference group |  |
| Stimulus spending in past 7 days | 1.464 | 1.291 - 1.661 | 1.553 | 1.444 - 1.671 |
| Household member lost work during the COVID-19 pandemic |  |  |  |  |
| Did not lose work | Reference group |  | Reference group |  |
| Lost work | 2.657 | 2.385 - 2.961 | 3.793 | 3.274 - 4.396 |
| Unemployment insurance |  |  |  |  |
| No unemployment insurance | Reference group |  | Reference group |  |
| Unemployment insurance | 0.894 | 0.766 - 1.044 | 1.090 | 0.968 - 1.228 |
| Any work in the past 7 days |  |  |  |  |
| Did not work in the past 7 days | Reference group |  | Reference group |  |
| Worked in the past 7 days | 0.845 | 0.593 - 1.203 | 0.567 | 0.417 - 0.771 |
| Wave and date |  |  |  |  |
| Wave 13, 8/19 to 8/31 | Reference group |  | Reference group |  |
| Wave 14, 9/2 to 9/14 | 1.000 | 0.853 - 1.173 | 0.965 | 0.735 - 1.268 |
| Wave 15, 9/16 to 9/28 | 1.025 | 0.839 - 1.251 | 0.996 | 0.856 - 1.158 |
| Wave 16, 9/30 to 10/12 | 1.165 | 0.915 - 1.482 | 1.056 | 0.853 - 1.307 |
| Wave 17, 10/14 to 10/26 | 1.183 | 0.977 - 1.432 | 1.144 | 0.958 - 1.366 |
| Wave 18, 10/28 to 11/9 | 1.157 | 0.913 - 1.466 | 1.106 | 0.916 - 1.334 |
| Wave 19, 11/11 to 11/23 | 1.274 | 1.013 - 1.602 | 1.345 | 1.044 - 1.733 |
| Wave 20, 11/25 to 12/7 | 1.196 | 0.985 - 1.452 | 1.315 | 1.036 - 1.668 |
| Wave 21, 12/9 to 12/21 | 1.336 | 1.037 - 1.721 | 1.278 | 0.968 - 1.687 |

**Appendix Table 5:**
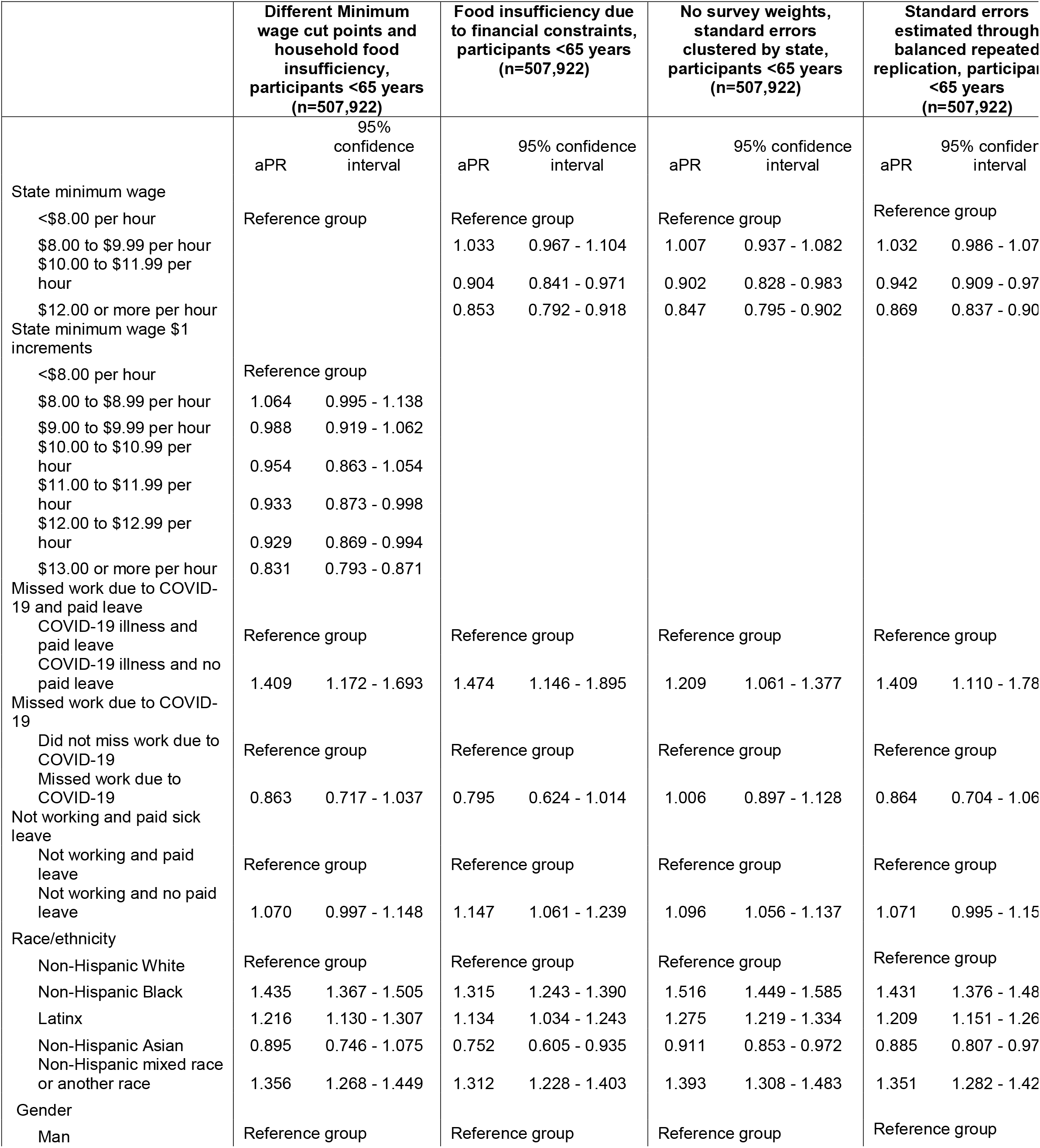
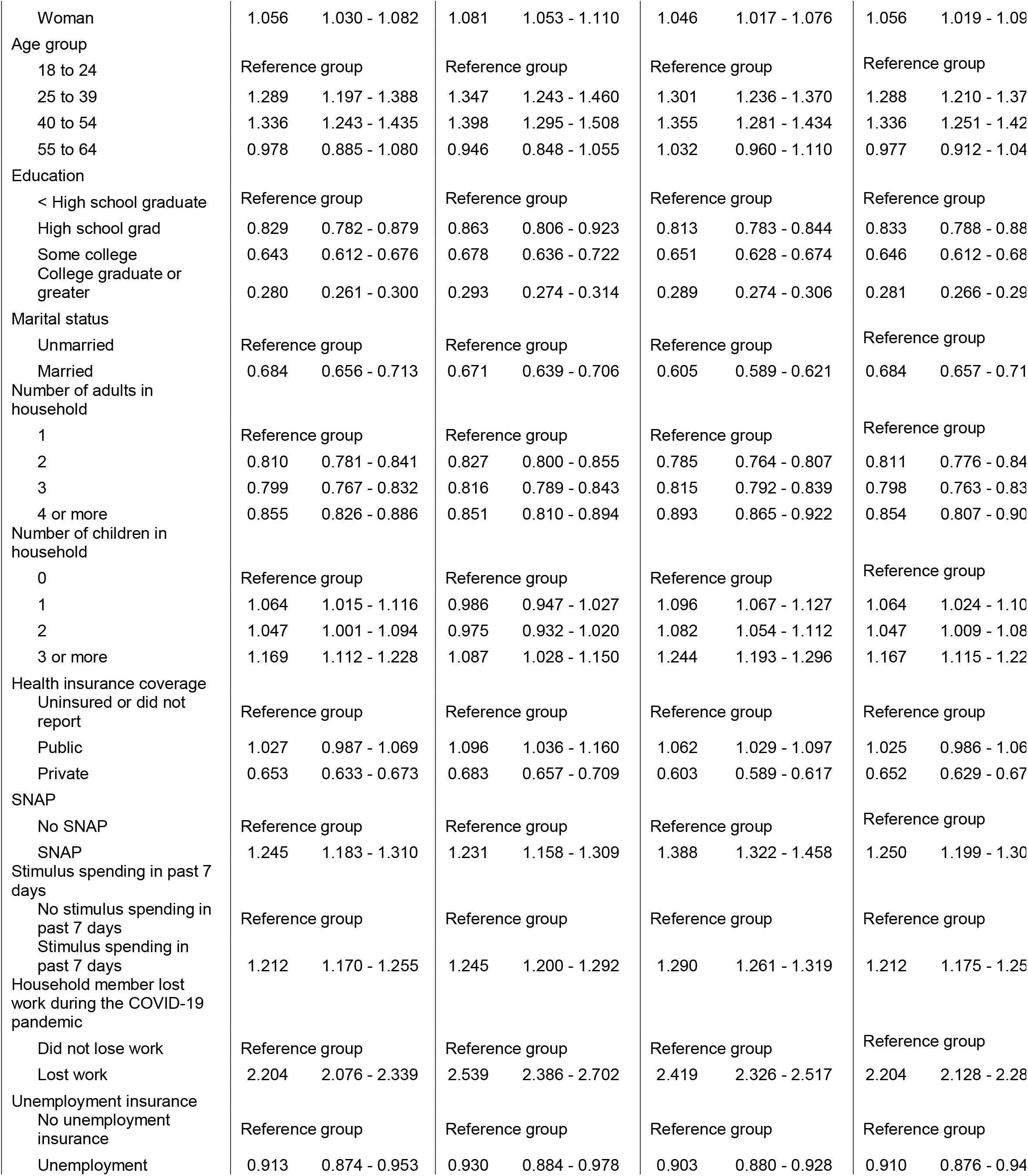
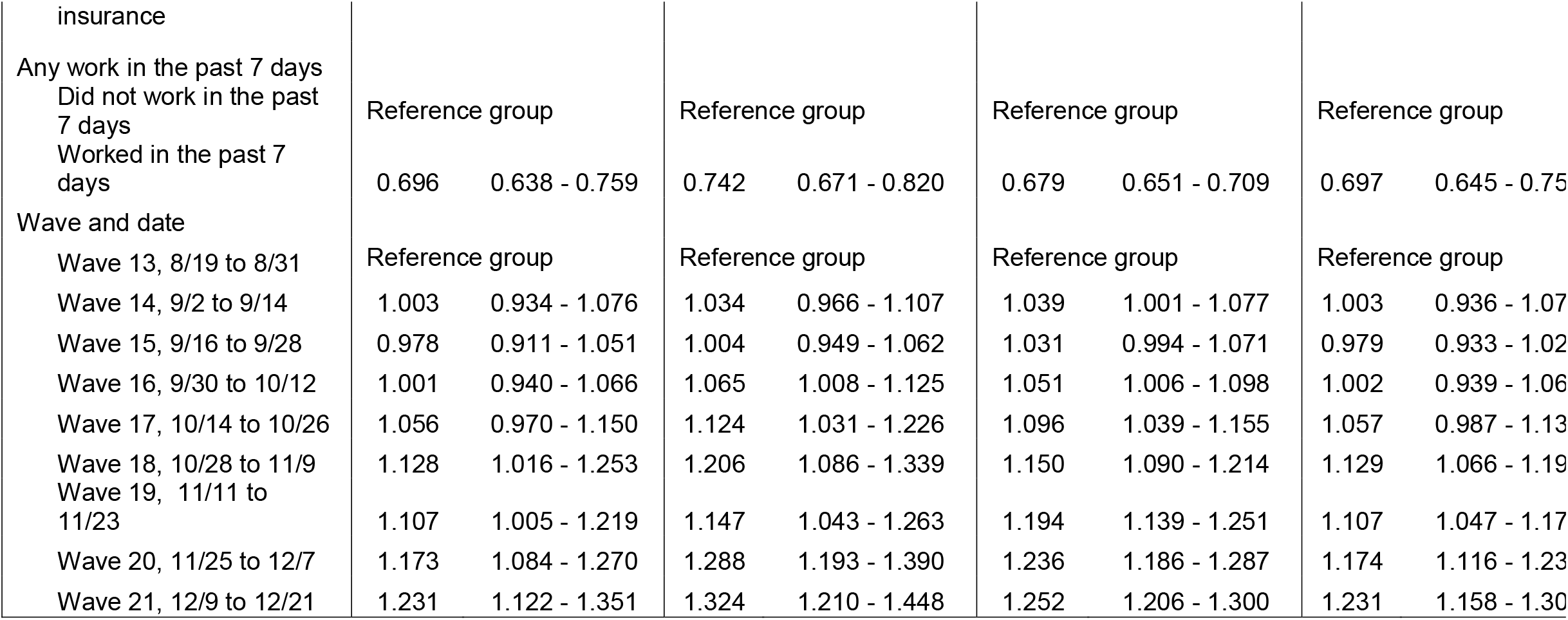
Sensitivity analyses state minimum wage policies and food insufficiency.

